# Psychiatric Comorbidities Predict Elevated Suicidal Symptoms in Children with ADHD

**DOI:** 10.1101/2025.08.09.25333367

**Authors:** Giulia Demo, Yi Xiong, Maria Ferrara, Jamal B. Williams

## Abstract

**Introduction:** Suicide in childhood is a growing clinical concern, and Attention-Deficit/Hyperactivity Disorder (ADHD) is increasingly recognized as a potential contributor. However, the role of psychiatric comorbidities, particularly depression, anxiety, and bipolar disorders, in shaping suicide risk in children with ADHD remains insufficiently understood

**Methods:** We analyzed data from 10,862 children aged 8–11 in the ABCD Study. Descriptive analyses were used to estimate the prevalence of suicidal behaviors and psychiatric comorbidities in children with and without ADHD, stratified by sex. Logistic regression models were applied to assess suicide risk associated with ADHD and comorbid psychiatric diagnoses, adjusting for age and sex.

**Results:** Children with ADHD had significantly higher rates of suicidal behaviors than controls. Passive suicidal ideation was reported in 15.6 % of females and 7.6 % of males with ADHD, versus 7.0 % and 4.7 % in controls (p < 0.001). Logistic regression identified Major Depressive Disorder (OR≈10–19) and Bipolar Disorder Type 2 (OR≈7–13) as the strongest predictors across outcomes. ADHD independently increased risk for most behaviors but moderated risk when comorbid with depression or Generalized Anxiety Disorder. Sex differences showed greater Bipolar Disorder Type 2- and Generalized Anxiety Disorder-related risk in females, and greater Bipolar Disorder Type 1- and depression-related risk in males.

**Discussion:** Children with ADHD have higher rates of suicidal thoughts and behaviors than controls, with Major Depressive Disorder and Bipolar Disorder Type 2 as the strongest independent predictors. However, ADHD moderates the risk from Major Depressive Disorder and General Anxiety Disorder, likely due to earlier recognition and intervention. Lastly, sex-specific patterns show higher BD2- and GAD-related risk in females, BD1-related risk in males.

**Conclusion:** ADHD significantly increases the risk of suicidal ideation in children. However, when present with comorbid conditions, ADHD may offer a clinical advantage by prompting earlier diagnosis and access to care. Improving ADHD recognition in pediatric settings could serve as a critical step in suicide prevention and early psychiatric intervention.

**PLAIN LANGUAGE SUMMARY:** Our study looked at more than 10,000 children aged 8–11 to see how ADHD and other mental health conditions affect suicidal thoughts and behaviors. Children with ADHD were more likely to have suicidal thoughts and behaviors than those without ADHD. Depression and Bipolar Disorder Type 2 were the strongest risk factors. Interestingly, when ADHD occurred alongside depression or Generalized Anxiety Disorder, the risk was often lower, possibly because ADHD leads to earlier detection and treatment. Risk patterns also differed by sex, suggesting that suicide prevention should be tailored to each child.

## INTRODUCTION

Suicide is a leading cause of death among youth in the United States, with an increasing rate among younger children ^1^. While puberty has long been considered the critical period for the onset of suicidal ideation, recent trends suggest that suicidal behaviors are emerging earlier, in prepubescent populations ^2^. National data show that suicide rose from the 10th to the 5th leading cause of death in children aged 5–11 between 2008 and 2019 ^3–5^. A cross-sectional analysis of U.S. emergency department data reported a 61% increase in visits for suicidal behavior between 2007 and 2015, with over 43% of these involving children aged 5 to 11 ^6^.

Multiple risk factors for suicide in children have been identified, including a history of suicide attempts, psychiatric diagnoses, and psychosocial adversity ^7^. One in four children who died by suicide had previously received care for a suicide attempt ^8^, and prior suicidal ideation is among the strongest predictors of future suicide risk ^9^. Sociodemographic factors such as race, gender, and access to care also influence risk, with higher rates seen among non-Hispanic white children and uninsured pre-adolescents ^7^. Exposure to abuse, trauma, or a family history of suicide further compounds vulnerability ^7^.

Psychiatric comorbidities, particularly mood and anxiety disorders, are major contributors to suicide risk ^10,11^. Approximately 80% of individuals who attempt suicide have a psychiatric diagnosis ^12^, and 89% of adolescents with suicidal behavior meet criteria for at least one disorder, including depression, anxiety, ADHD, and oppositional defiant disorder ^13^. Children with psychiatric diagnoses have suicide attempt rates up to five times higher than their peers ^3^. ADHD is the most common psychological disorder in school-aged children ^14,15^, with U.S. prevalence estimates ranging from 3% to 11% ^16^. Its frequent comorbidity with depression and anxiety contributes to significant emotional dysregulation and functional impairment ^17,18^.

Children with both ADHD and depression experience more severe symptoms than those with either condition alone ^19^. Additionally, the overlap between ADHD and Bipolar Disorder (BD) is clinically relevant: about 85% of children with BD also meet criteria for ADHD, and 22% of children with ADHD are diagnosed with BD ^20^. The lifetime suicide risk in individuals with BD is at least 15 times higher than in the general population ^21^.

Despite these associations, suicidality in young children, particularly those with ADHD, remains underexplored, partly due to misconceptions about their ability to comprehend or act on suicidal thoughts ^22^. Yet children as young as seven can understand the permanence and universality of death ^23^, suggesting that early suicidal behavior may reflect intentional ideation.

This study aims to investigate how psychiatric comorbidities, specifically depression, anxiety, and BD, influence the risk of suicidal ideation in children with and without ADHD. Using data from the ABCD Study, we explore whether ADHD amplifies or modifies the impact of these comorbidities on suicidality. Specifically, we ask if children with ADHD exhibit a higher risk of suicidal behaviors compared to their peers with psychiatric diagnoses and no ADHD. And which psychiatric comorbidities are most strongly associated with suicidal ideation in children with ADHD?

## METHODS

### Data and Participant Selection

This study utilized data from Release 5.1 of the Adolescent Brain Cognitive Development (ABCD) Study, a large, longitudinal dataset collected from baseline through two-year follow-up assessments. Our sample consisted of 10,862 children, aged between 8.91 years (107 months) and 11.08 years (133 months) (**Supplemental Table 1**), selected based on confirmed clinical diagnoses of ADHD, psychiatric conditions, or suicidal behaviors ^24^. The psychiatric diagnosis was assessed using the Schedule for Affective Disorders and Schizophrenia for School-Age Children, Present and Lifetime Version (K-SADS-501) diagnostic tool. This semi-structured interview, based on DSM-5 diagnostic criteria and administered by trained clinical interviewers, is designed for children aged 6–17 years and evaluates past, current, and partially remitted episodes of psychopathology ^25^. Each diagnostic variable was recorded as a binary value, with 0 indicating the absence of the diagnosis and 1 indicating its presence.

### Measurements and Key Variables

Suicidal ideation exists along a spectrum of intensity, beginning with a general desire to die, which may lack specific methods, plans, or concrete intentions, and extending to the actual execution, planning, and preparation for a definitive attempt to end one’s life ^26,27^. To streamline our dataset and reduce the number of variables, we consolidated several related measures into binary groups. Specifically, we created a single variable for “Active Suicidal Ideation,” which takes the value of 1 if a patient exhibits at least one of the following behaviors: Active Intent, Active Methods, Active Non-Specific Ideations, or Active Plan, and 0 if none are present. We then did the same thing for “Aborted or Interrupted Attempt”. Furthermore, we combined both current and past ideation data into unified binary variables, ultimately yielding five binary variables to capture suicidality, which we call Suicidal Behaviors: Aborted or Interrupted Attempt, Active Suicidal Ideation, Passive Suicidal Ideation, Preparatory Actions Toward Imminent Suicidal Behavior, and Suicide Attempt (**Supplemental Table 2**).

Similarly to the approach used for the suicidal behaviors, we consolidated the psychiatric comorbidity measures into eight binary variables. These represent: Bipolar Disorder Type 1 (BD1), Bipolar Disorder Type 2 (BD2), Dysthymia, Generalized Anxiety Disorder (GAD), Major Depressive Disorder (MDD), Social Anxiety Disorder (SAD), Unspecified Bipolar and Related Disorder (UBD), and Unspecified Depressive Disorder (UDD) (**Supplemental Table 3**). Each variable was coded as 1 if the patient met the diagnostic criteria for that condition, and 0 if not.

#### Statistical Analysis

### Descriptive Analysis

Bivariate descriptive analyses were conducted to estimate the frequency of suicidal behaviors and their associated risk factors within the sample, as well as to examine sex differences between children with and without an ADHD diagnosis. Prevalence rates were calculated and compared using chi-square tests for categorical comparisons, with statistical significance set at p < 0.05.

The distribution of psychiatric disorders linked to suicidal risk was also analyzed to assess their relative impact within ADHD and control groups.

### Multivariate Logistic Regression Analysis

Multivariate logistic regression models were used to quantify the associations between ADHD status, psychiatric comorbidities, and suicidal behaviors, controlling for age and sex. Separate logistic regression models were fitted individually for each suicidal behavior (Aborted or Interrupted Suicide Attempt, Passive Suicidal Ideation, Active Suicidal Ideation, Preparatory Actions Toward Imminent Suicidal Behavior, and Suicide Attempt). The general logistic regression model structure was as follows:

General Model:

𝑙𝑜𝑔𝑖𝑡(𝑃(𝑠𝑢𝑖𝑐𝑖𝑑𝑎𝑙 𝑏𝑒ℎ𝑎𝑣𝑖𝑜𝑟))

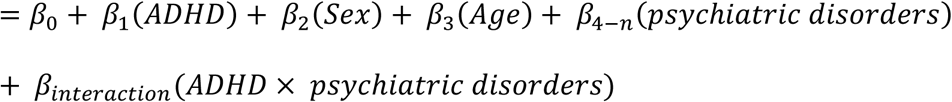

To identify sex-dependent differences in ADHD-related suicide risk, logistic regression analyses were performed separately by sex. The sex differences in Odds Ratios (OR) were calculated by subtracting OR Male from OR Female. Positive values represent relatively higher OR for females, and negative values indicate relatively higher OR in males. The sex-stratified logistic regression model was structured as follows:

Sex-Stratified Model:

𝑙𝑜𝑔𝑖𝑡(𝑃(𝑠𝑢𝑖𝑐𝑖𝑑𝑎𝑙 𝑏𝑒ℎ𝑎𝑣𝑖𝑜𝑟))

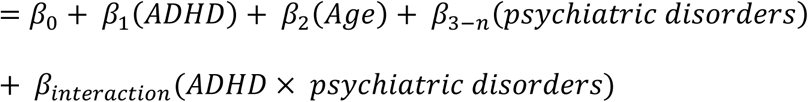

OR and 95% confidence intervals were calculated by exponentiating coefficients of the logistic regression. Predictors were then evaluated using Wald Z-tests, with significance assigned using the convention *p < 0.05, **p < 0.01, ***p < 0.001. Additionally, these models were fitted using Python’s statsmodels version 0.14.0.

### Visualization of Effect Sizes

Finally, to evaluate the strength and direction of these associations, Z-values were derived from the regression analysis and visualized as a heatmap. Positive Z-values indicate increased risk, negative values represent decreased risk, and intensity reflects the magnitude of the association.

Data Stability and Exclusion Criteria

For the most robust and reliable results, subsets containing fewer than 20 participants or lacking sufficient variability were excluded from logistic regression analyses.

## RESULTS

### Phase 1: Descriptive Analysis of the Study Cohort

We analyzed data from 10,862 children aged 8–11 years (107–133 months), including 5,664 males and 5,198 females. Of these, 1,751 were diagnosed with ADHD (1,192 males, 559 females), and 9,111 served as controls (4,472 males, 4,639 females) (**Supplemental Table 1**). Ancestry distribution included 2,107 children of African descent, 2,074 Native American, 6,551 European, 75 South Asian, and 55 East Asian (**Supplemental Table 1**).

Children diagnosed with ADHD showed significantly higher prevalence rates of suicidal behaviors compared to controls (**Table 1**). Specifically, females with ADHD exhibited notably higher rates of passive suicidal ideation (15.6% ADHD vs. 7.0% controls, p < 0.001), active suicidal ideation (8.9% ADHD vs. 5.0% controls, p < 0.001), and suicide attempts (3.0% ADHD vs. 1.3% controls, p < 0.01) (**Fig. 1a**). Male children with ADHD also had significantly elevated prevalence of passive suicidal ideation (7.6% ADHD vs. 4.7% controls, p < 0.001), active suicidal ideation (6.0% ADHD vs. 3.4% controls, p < 0.001), and suicide attempts (2.2% ADHD vs. 0.7% controls, p < 0.001) (**Fig. 1a**). Prevalence of aborted or interrupted attempts and preparatory actions toward imminent suicidal behavior was also consistently higher in the ADHD group, particularly among females (**Table 1**), though differences in these behaviors were only significant in females (2.5% ADHD vs. 0.6% controls, p < 0.001) (**Fig. 1a**). These results highlight that ADHD significantly elevates the risk of suicidal behaviors in children, with females generally demonstrating higher absolute prevalence rates compared to males.

**Figure 1.**
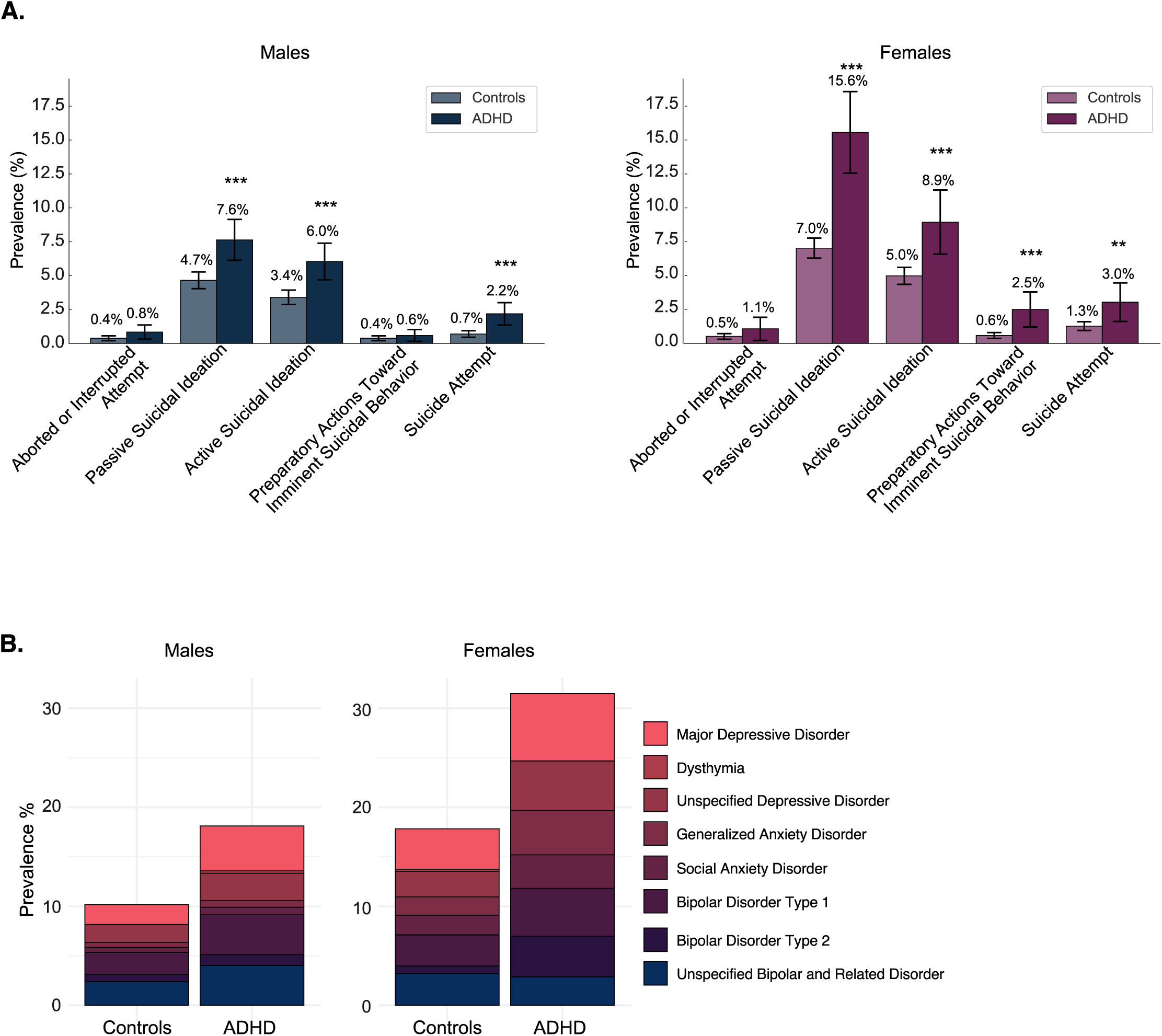
Prevalence of Suicidal Behaviors and Psychiatric Comorbidities Among Children with ADHD. (A) Prevalence (%) of suicidal behaviors among male and female children with ADHD compared to neurotypical controls. Significant differences indicated by asterisks (*p < 0.05, **p < 0.01, ***p < 0.001). Error bars represent the 95% confidence intervals. (B) Stacked bar charts of the prevalence (%) of psychiatric comorbidities stratified by sex and ADHD status.

**Table 1.** Prevalence of suicidal behaviors in children with and without ADHD, stratified by sex. Prevalence % of Passive Suicidal Ideation, Active Suicidal Ideation, Suicide Attempt, Preparatory Actions Toward Imminent Suicidal Behavior, and Aborted or Interrupted Attempt in the study cohort.

| Sex | Behavior | ADHD<br>Prevalence<br>(%) | Chi-square | P-value |
| --- | --- | --- | --- | --- |
| Male | Aborted or Interrupted Attempt | 0.83892617<br>4 | 3.26441572<br>2 | 0.07079823<br>9 |
| Male | Passive Suicidal Ideation | 7.63422818<br>8 | 16.1576189<br>7 | 5.83E-05 |
| Male | Active Suicidal Ideation | 6.04026845<br>6 | 16.5979714<br>7 | 4.62E-05 |
| Male | Preparatory Actions toward imminent<br>Suicidal behavior | 0.58724832<br>2 | 0.52884639 | 0.46709226<br>5 |
| Male | Suicide Attempt | 2.18120805<br>4 | 19.4503039<br>8 | 1.03E-05 |
| Female | Aborted or Interrupted Attempt | 1.07334525<br>9 | 1.80600022<br>6 | 0.17898878<br>6 |
| Female | Passive Suicidal Ideation | 15.5635062<br>6 | 48.5404647<br>8 | 3.24E-12 |
| Female | Active Suicidal Ideation | 8.94454382<br>8 | 14.5720179<br>3 | 0.00013490<br>3 |
| Female | Preparatory Actions toward imminent<br>Suicidal behavior | 2.50447227<br>2 | 21.1688010<br>1 | 4.21E-06 |
| Female | Suicide Attempt | 3.04114490<br>2 | 9.64676363<br>7 | 0.00189685<br>5 |

Children diagnosed with ADHD had significantly higher prevalence rates of psychiatric conditions compared to controls (**Table 2**). Females with ADHD showed notably higher prevalence rates of MDD (6.8% ADHD vs. 4.1% controls), BD2 (4.1% ADHD vs. 0.8% controls), GAD (4.5% ADHD vs. 1.9% controls), and SAD (3.4% ADHD vs. 2.0% controls) (**Fig. 1b**). Similarly, males with ADHD demonstrated elevated prevalence of MDD (4.5% ADHD vs. 2.0% controls), BD1 (4.0% ADHD vs. 2.2% controls), and UDD (2.8% ADHD vs. 1.8% controls) (**Fig. 1b**). Across psychiatric diagnoses, females typically show higher overall prevalence rates compared to males, suggesting potential sex-dependent differences in psychiatric burden associated with ADHD.

**Table 2.** Prevalence of psychiatric comorbidities in children with and without ADHD, stratified by sex. Prevalence of psychiatric conditions in the study cohort.

| Sex | Trait | Unspecified Bipolar and Related Disorder | Bipolar Disorder Type 2 | Bipolar Disorder Type 1 | Social Anxiety Disorder | Generalized Anxiety Disorder | Unspecified Depressive Disorder | Dysthymia | Major Depressive Disorder |
| --- | --- | --- | --- | --- | --- | --- | --- | --- | --- |
| Male | Controls | 2.39266<br>5474 | 0.71556<br>3506 | 2.23613<br>5957 | 0.49194<br>9911 | 0.5143<br>1127 | 1.8112<br>7013 | 0 | 2.01252<br>2361 |
| Male | ADHD | 4.02684<br>5638 | 1.09060<br>4027 | 4.02684<br>5638 | 0.75503<br>3557 | 0.6711<br>4094 | 2.7684<br>5638 | 0.2516<br>7785 | 4.53020<br>1342 |
| Female | Controls | 3.21189<br>9116 | 0.75447<br>2947 | 3.14723<br>0006 | 1.98318<br>6031 | 1.8538<br>4781 | 2.5867<br>6439 | 0.2155<br>637 | 4.07415<br>3912 |
| Female | ADHD | 2.86225<br>4025 | 4.11449<br>0161 | 4.83005<br>3667 | 3.39892<br>6655 | 4.4722<br>7191 | 5.0089<br>4454 | 0 | 6.79785<br>3309 |

### Phase 2: Multivariate Analysis of ADHD, Psychiatric Conditions, and Interaction Effects on Suicidal Risk

For each suicidal outcome, multivariate logistic regression was used to access the independent contributions of ADHD, psychiatric conditions, and ADHD × comorbidity interactions (**Fig. 2**; **Supplementary** Fig.1). For *Suicidal Attempt*, MDD (OR ≈ 19.4, 95 % CI ≈ 11.6–32.3) was the strongest independent predictor, with BD2 (OR ≈ 6.8, CI ≈ 2.5–18.3) and BD1 (OR ≈ 4.6, CI ≈ 2.2–9.7) also significantly increasing risk (**Fig. 2a**). ADHD was also a significant risk factor (OR ≈ 2.9, CI ≈ 1.8–4.8) (**Fig. 2a**), indicating that children with ADHD are nearly three times more likely to attempt suicide than controls. However, significant interaction between ADHD and MDD (OR ≈ 0.36, CI ≈ 0.14–0.91) suggested that while depression elevates risk substantially (**Fig. 2a**), this effect is less pronounced in children with ADHD than in those without ADHD. *Preparatory actions toward imminent suicidal behavior* was most strongly impacted by MDD (OR ≈ 16.7, CI ≈ 8.1–34.7) and BD2 (OR ≈ 13.3, CI ≈ 4.0–44.5) (**Fig. 2b**). GAD (OR ≈ 4.5, CI ≈ 1.8–11.1) and ADHD (OR ≈ 3.7, CI ≈ 1.8–7.4) also increased risk (**Fig. 2b**). Again, the ADHD and MDD interaction (OR ≈ 0.03, CI ≈ 0.00–0.30) indicated that the heightened risk associated with depression was dramatically attenuated among children with ADHD (**Fig. 2b**). A considerable amount of psychiatric conditions were significant predictors of *Active suicidal ideation* including, MDD (OR ≈ 12.8, CI ≈ 9.5–17.4), BD2 (OR ≈ 7.4, CI ≈ 4.0–13.6), UDD (OR ≈ 3.1, CI ≈ 2.1–4.8), GAD (OR ≈ 3.7, CI ≈ 2.2–6.2), BD1 (OR ≈ 2.5, CI ≈ 1.6–4.0), and SAD (OR ≈ 2.3, CI ≈ 1.4–3.7) (**Fig. 2c**). ADHD alone remained an independent risk factor (OR ≈ 1.7, CI ≈ 1.3–2.3) (**Fig. 2c**). Significant interactions showed that ADHD moderated the risk conferred by MDD (OR ≈ 0.45, CI ≈ 0.24–0.82) and GAD (OR ≈ 0.25, CI ≈ 0.08–0.76) (**Fig. 2c**). Additionally, the interaction of ADHD with unspecified bipolar disorder (OR ≈ 2.54, CI ≈ 1.09–5.92) suggested that having ADHD amplified the risk in children with UBD (**Fig. 2c**).

**Figure 2.**
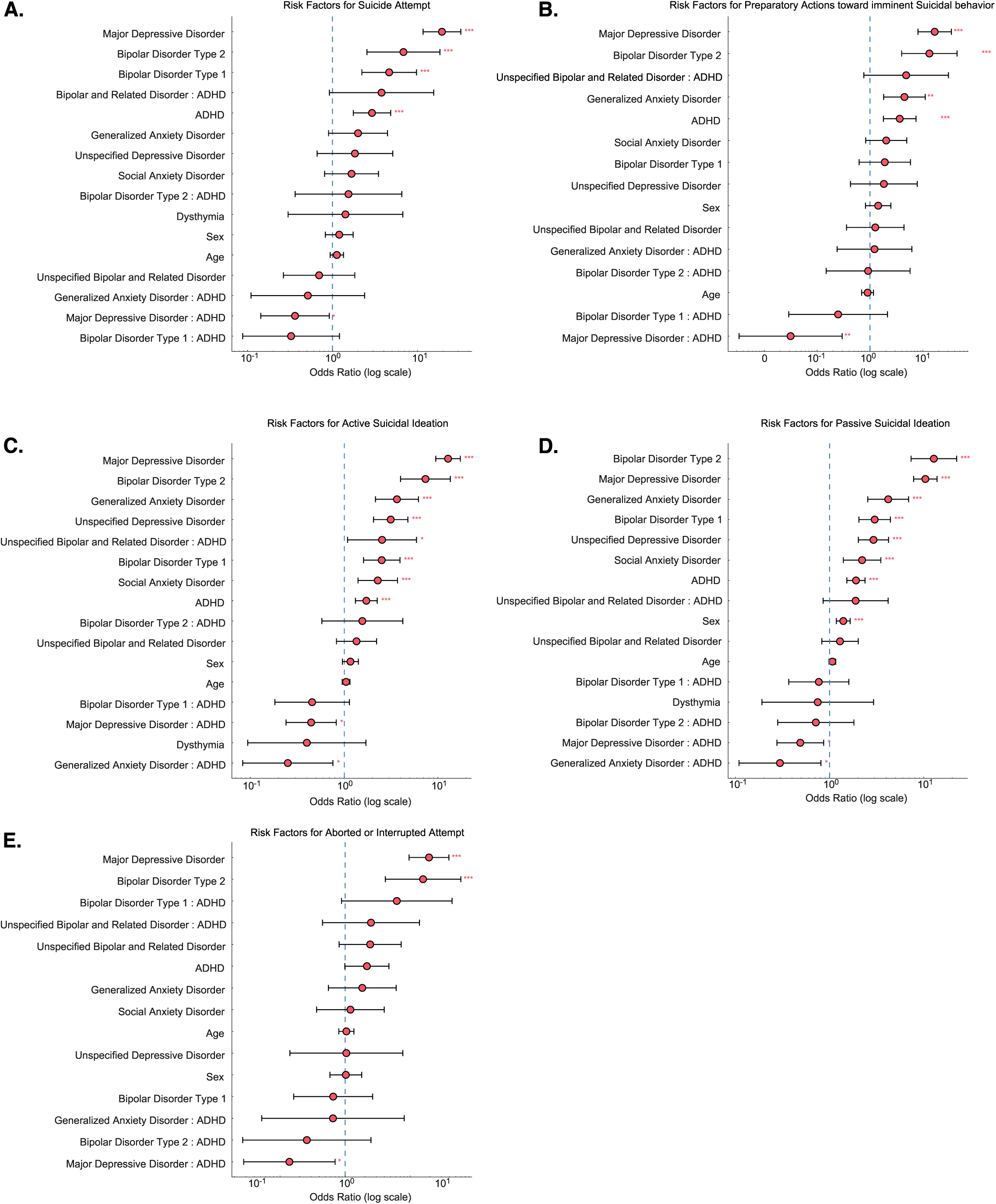
Psychiatric Risk Factors Associated with Suicidal Behaviors in Children with ADHD. Forest plots of odds ratios with 95% confidence intervals for risk factors associated with suicidal behaviors (A) Suicide Attempts, (B) Preparatory Actions toward Imminent Suicidal Behavior, (C) Active Suicidal Ideation, (D) Passive Suicidal Ideation, (E) Aborted or Interrupted Suicide Attempts. Significant associations marked by asterisks (*p < 0.05, **p < 0.01, ***p < 0.001). Dashed line indicates no effect (OR = 1).

For *Passive suicidal ideation*, BD2 (OR ≈ 12.8, CI ≈ 7.3–22.3) and MDD (OR ≈ 10.4, CI ≈ 7.8–13.8) showed the strongest associations (**Fig. 2d**). GAD (OR ≈ 4.2, CI ≈ 2.5–6.9), BD1 (OR ≈ 3.0, CI ≈ 2.1–4.4), UDD (OR ≈ 2.9, CI ≈ 2.0–4.2), SAD (OR ≈ 2.2, CI ≈ 1.4–3.5), and ADHD (OR ≈ 1.9, CI ≈ 1.5–2.4) were also significant (**Fig. 2d**). Sex also modestly increased risk (OR ≈ 1.4, CI ≈ 1.2–1.7) (**Fig. 2d**). ADHD significantly moderated risk associated with MDD (interaction OR ≈ 0.49, CI ≈ 0.28–0.87) and GAD (interaction OR ≈ 0.30, CI ≈ 0.11–0.81) (**Fig. 2d**), indicating lower incremental risk for these disorders among children with ADHD. Lastly, *Aborted or interrupted suicide attempts* showed that MDD (OR ≈ 19.4, CI ≈ 9.6–39.3) and BD2 (OR ≈ 15.7, CI ≈ 4.1–59.8) were the only significantly associated predictors (**Fig. 2e**). However, ADHD alone was not a significant predictor. Though, the ADHD and MDD interaction (OR ≈ 0.14, CI ≈ 0.03–0.70) was again significant (**Fig. 2e**), indicating that the elevated risk of aborted attempts in depressed children was substantially lower in those with ADHD than in those without ADHD.

Together, these data show that several psychiatric disorders, particularly MDD and BD2 are strongly associated with suicidal behaviors across outcomes. ADHD itself is an independent risk factor for suicide attempts, preparatory actions, active ideation, and passive ideation, but not for aborted or interrupted attempts. Also, ADHD consistently moderates (and in some cases mitigates) the added risk conferred by MDD and generalized anxiety disorder, showing that there is a complex relationship between ADHD and psychiatric comorbidities in shaping suicide risk among children.

### Phase 3: Sex Differences in the Association Between Psychiatric Diagnoses and Suicidal Behaviors

We then measured sex differences by comparing the OR between females and males for each combination of psychiatric diagnosis and suicidal behavior, controlling for ADHD (**Fig. 3**). Females with BD2 showed the most significant risk across multiple outcomes. In particular, females diagnosed with BD2 had much higher odds differences for *aborted or interrupted attempts* (difference = 3.40), *preparatory actions* (difference = 2.99), *suicide attempts* (difference = 2.50), and *passive ideation* (difference = 0.95) relative to males. Another considerable female-dominant effect appeared for females with UBDs in *preparatory actions* (difference = 3.30). Notably, females with GAD had a much greater risk of *aborted or interrupted attempts* (difference = 2.80) and *suicide attempts* (difference = 1.15) compared with males. Smaller but still positive differences were observed for BD2 in *active suicidal ideation* (difference = 0.79) and *passive suicidal ideation* (difference = 0.95). UBD also showed positive differences for *active suicidal ideation* (difference = 0.71) and *passive suicidal ideation* (difference = 1.69). Additionally, GAD in *active suicidal ideation* showed a positive difference (difference = 0.57).

**Figure 3.**
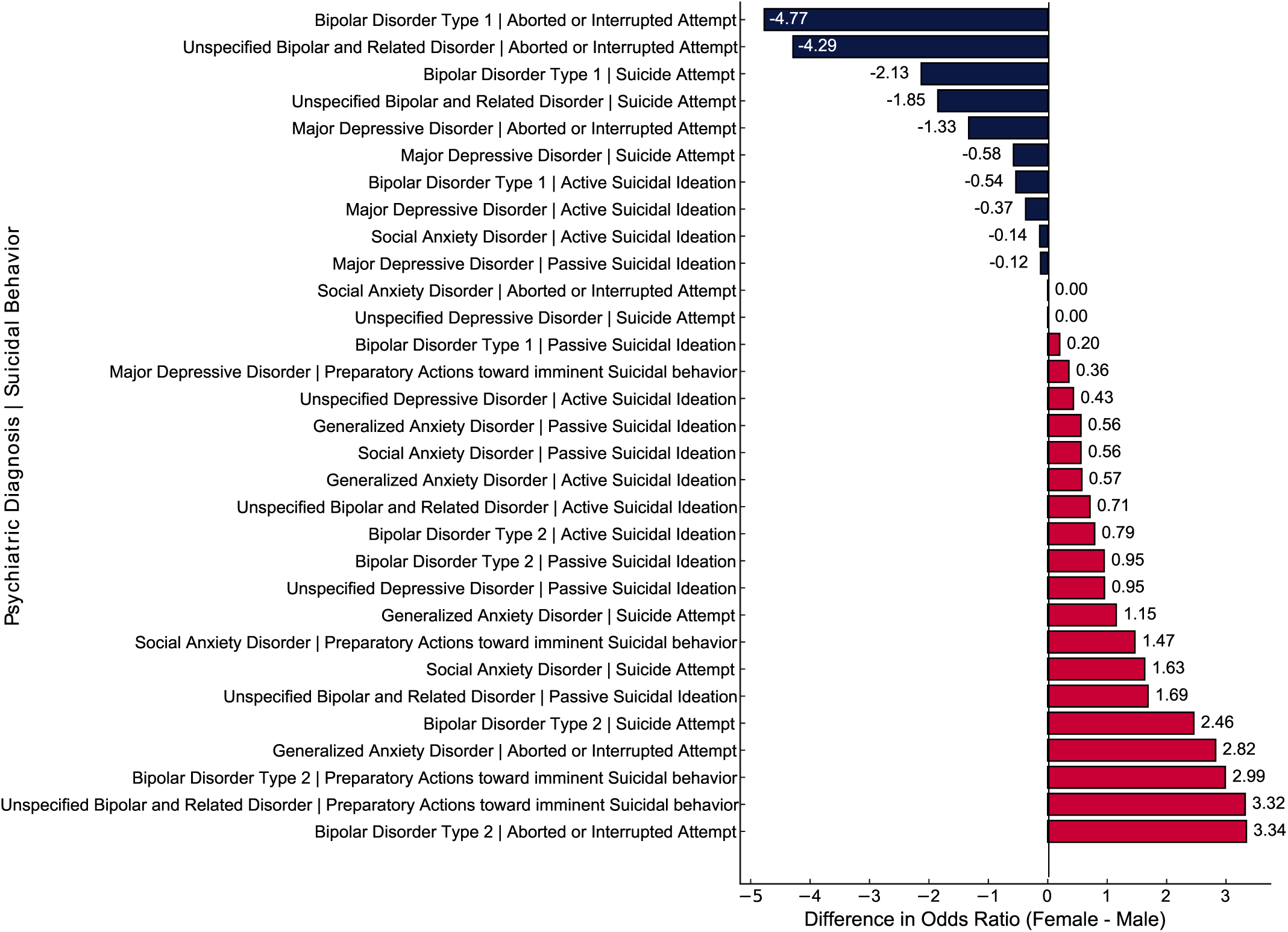
Sex Differences in ADHD-Related Suicide Risk. Bar graph of differences in odds ratios between females and males across psychiatric diagnoses and suicidal behaviors. Positive values (red) indicate a higher odds ratio in females; negative values (blue) indicate higher odds in males.

Males exhibited a greater risk in several areas, including BD1, which was associated with a much higher risk of *aborted or interrupted attempts* than in females (difference = –4.77), and this male-predominant pattern persisted for UBD (difference = –4.29). For *suicide attempts*, males with BD1 and UBD remained at higher risk (differences = –2.13 and –1.85, respectively). MDD also showed a moderately higher risk of *aborted or interrupted attempts* (difference = – 1.33) and *suicide attempts* (difference = –0.58) in males. Smaller male-associated differences were noted for *active ideation* linked to BD1 (difference = –0.54) and MDD (difference = – 0.37). Lastly, several combinations showed differences near zero, indicating similar risks in males and females.

## DISCUSSION

### ADHD and Early Suicidal Ideation Represent a Blind Spot in Pediatric Mental Health

Research on suicide in preschool-aged children remains limited, with most of the attention focused on puberty as the critical period for the emergence of suicidal ideation. However, numerous studies have shown that even very young children can experience suicidal thoughts. Why, then, is this population so consistently underestimated? A widespread belief persists that preschool-aged children are not cognitively capable of processing the concept of death or forming an intention to die.

Yet suicide exists along a continuum of thoughts and behaviors, not solely in its fatal outcomes. As noted by Wasserman et al. (2021), the risk factors for suicide in children and adolescents often mirror those seen in adults. Importantly, one of the strongest predictors of a future suicide attempt is a past suicidal thought. This implies that children who express suicidal ideation at an early age already represent a high-risk group and require early and sustained attention. A common misconception is that children cannot truly grasp what death means. However, Speece and Brent (1984) showed that by around age seven, most children begin to understand death as permanent, universal, and resulting in the cessation of function ^23^. Once this developmental milestone is reached, if a child begins to exhibit self-harming or suicidal behaviors, any act related to death, whether completed or not, can no longer be dismissed as unintentional. Instead, such actions must be understood as part of the broader spectrum of suicide and intentional end-of-life behavior.

Our study contributes to this body of knowledge by demonstrating a strong correlation between ADHD and suicidal behaviors in children. Given that ADHD diagnoses are rising globally, this population represents not only a clinically relevant group but also a rapidly expanding one. Our Phase 1 analyses reinforce this urgency. Passive suicidal ideation was the most common behavior, but active ideation, preparatory actions, and attempts also occurred. Across all behaviors, prevalence was higher in children with ADHD than in controls. For instance, 15.6 % of females with ADHD and 7.6 % of males with ADHD reported passive ideation, compared with 7.0 % and 4.7 % in female and male controls, respectively. Active ideation and attempts showed similar trends. These findings highlight that ADHD identifies a growing pediatric subpopulation at distinctly elevated risk for suicidal thoughts and behaviors, emphasizing the need for early recognition.

### Risk and Resilience within ADHD, Psychiatric Comorbidities, and Suicidal Behaviors

ADHD itself was an independent risk factor for several suicidal behaviors. In Phase 2 we show that ADHD increased the odds of active and passive ideation, preparatory actions and suicide attempts by factors of about 1.7–2.9 after controlling for covariates. Among psychiatric conditions, MDD and BD2 were the strongest predictors across outcomes. MDD was linked to a very high risk of suicide attempts and both forms of ideation, whereas BD2 was connected to a high risk of ideation and preparatory behaviors and a moderate risk of attempts. GAD and SAD were also significant for certain behaviors.

Remarkably, ADHD moderated several of these associations. Interaction terms showed that children with ADHD had lower incremental risk when MDD or GAD was present, suggesting that an ADHD diagnosis may trigger earlier recognition of psychiatric symptoms and interventions that dampen risk. ADHD did not, however, mitigate the risk associated with BD2 or BD1, indicating that mood disorders with pronounced episodic features may operate through distinct mechanisms. These nuanced patterns highlight the importance of considering both main effects and interactions when assessing suicide risk in children with complex psychiatric profiles.

### Sex-Specific Patterns of Suicidal Risk

Phase 3 analyses revealed notable sex differences in the association between psychiatric diagnoses and suicidal behaviors. These analyses show that certain diagnoses, like BD2, UBD, and GAD, carried a higher relative risk for specific suicidal behaviors in females, while BD1 and UBD carried a higher risk in males. These differences indicate the value of sex-specific risk assessment in clinical practice.

### From diagnosis to outcome: early recognition as an opportunity for resilience

One of the most striking findings is that, within children who had psychiatric comorbidities, those with ADHD often exhibited lower incremental risk than those without ADHD when comorbid MDD or GAD was present. This apparent contradiction may reflect earlier recognition and management triggered by ADHD symptoms. ADHD shares neurocognitive traits with suicidality, such as emotional dysregulation and impulsivity, that prompt clinicians and families to seek evaluation earlier than they might for internalizing symptoms alone ^1^. Because ADHD is socially accepted as a behavioral and academic issue, it may serve as a gateway to services.

Treatment of ADHD often improves emotional regulation, social functioning and coping skills, and these improvements may generalize to comorbid conditions. Our data also show that BD2 and GAD were the comorbidities most strongly linked to suicidality in children with ADHD, consistent with evidence that ADHD can evolve into mood disorders when additional risk factors exist ^28^. Early diagnosis and management of ADHD may therefore break this trajectory and reduce suicide risk.

### Translational Perspectives for Suicide Prevention in Children with Psychiatric Risk

These findings raise several translational implications. First, clinicians should actively screen for suicidal thoughts in children with ADHD and comorbid mood or anxiety disorders, including those younger than adolescence. Second, interventions should address both ADHD and comorbid conditions, such as programs enhancing executive function and emotional regulation that may reduce suicidality. Third, sex-specific risk profiles emphasize the need for tailored approaches. Females with BD2 or GAD and males with BD1 or unspecified bipolar disorder warrant targeted prevention efforts. Finally, stigma remains a barrier; destigmatizing mood and anxiety disorders in children and improving precision in diagnostic processes could help ensure timely, comprehensive care.

## CONCLUSION

This study identifies ADHD as both a marker of heightened suicide risk in children and, in certain contexts, a moderator that can attenuate the additional risk posed by specific comorbidities such as MDD and GAD. Early ADHD diagnosis may facilitate earlier recognition of psychiatric distress, increasing opportunities for timely intervention. However, the persistent and substantial risk linked to mood disorders with episodic features, particularly BD2, underscores the need for careful monitoring and targeted prevention. Incorporating ADHD screening into suicide prevention strategies, along with sex-specific risk assessment and intervention, could play a critical role in reducing childhood suicide risk.

## Data Availability

All data produced in the present study are available upon reasonable request to the authors

## ACKNOWLEDGEMENTS

This research used data from the Adolescent Brain Cognitive DevelopmentSM (ABCD) Study (https://abcdstudy.org), supported by the National Institutes of Health and other federal partners (awards U01DA041048, U01DA050989, U01DA051016, U01DA041022, U01DA051018, U01DA051037, U01DA050987, U01DA041174, U01DA041106, U01DA041117, U01DA041028, U01DA041134, U01DA050988, U01DA051039, U01DA041156, U01DA041025, and U01DA041120). A full list of supporters is available at https://abcdstudy.org/federal-partners.html, and consortium members are listed at https://abcdstudy.org/consortium_members/. The investigators within the ABCD consortium provided data but were not involved in analysis or manuscript preparation. This manuscript used ABCD data release 5.1. We thank Yana Shimanovich for her assistance in helping the team access the variables used in this study.

## CONTRIBUTIONS

G.D., M.F., and J.B.W. conceived and designed experiments. Y.X., M.F., and J.B.W. suggested analyses. M.F. and J.B.W. supervised the project. G.D. performed analyses. G.D. and J.B.W. wrote the manuscript. M.F. edited the manuscript.

## ETHICS DECLARATIONS

This study only utilized data from the ABCD Study release version 5.1, which was downloaded from the National Institute of Mental Health Data Archive ^29^. The dataset (v5.1) includes full cohort data from the baseline and the first two years of follow-up, which were the primary focus of the current investigation. The ABCD Study was approved by the institutional review board (IRB) at the University of California (central IRB). Informed consent for human research was obtained from both caregivers and participants. This current study is therefore exempt from informed consent by the University at Buffalo’s Institutional Review Board. The methods used in this study reveal no identifying information of the subjects.

---

**Supplemental Figure 1.**
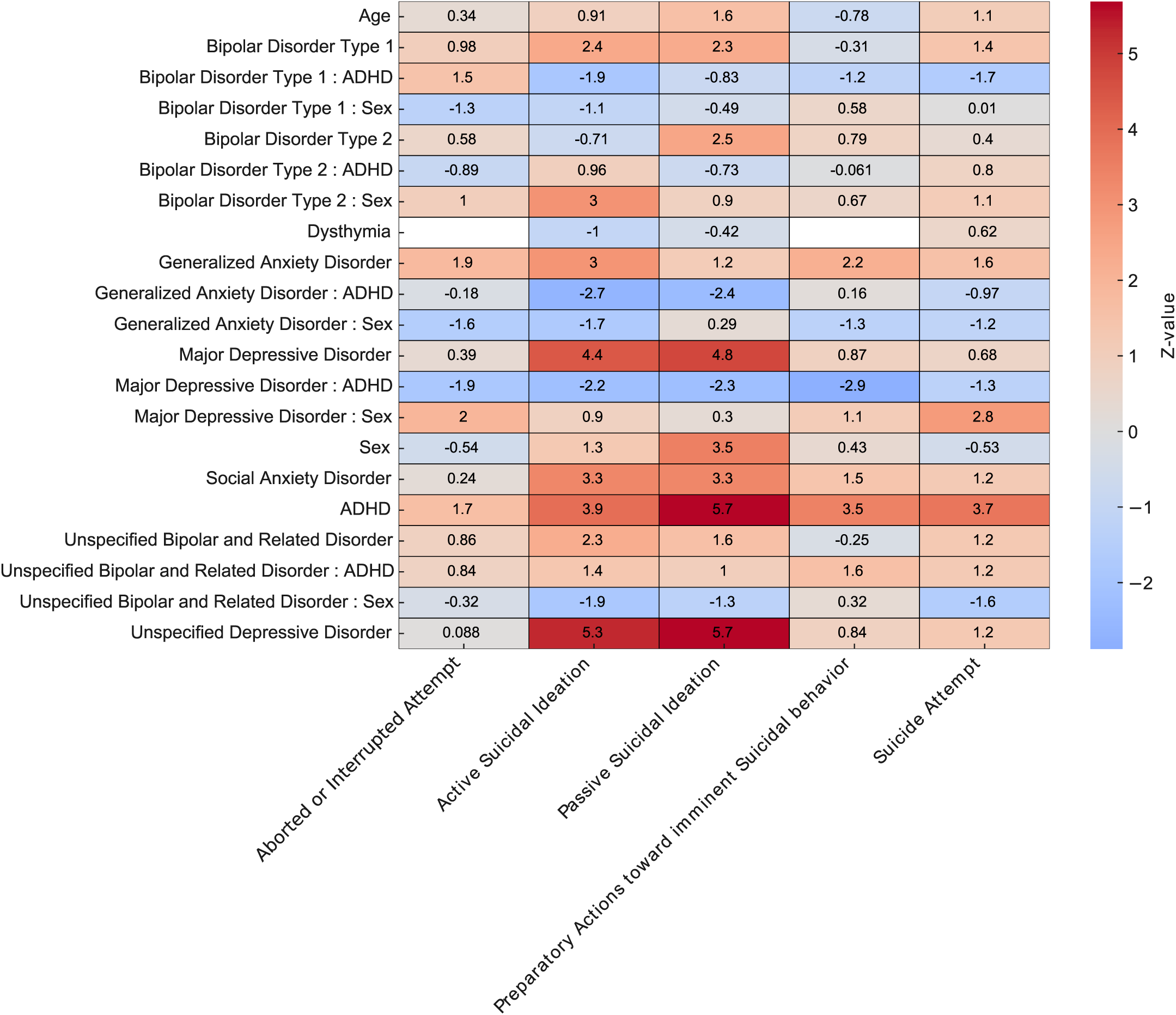
Heatmap of Effect Sizes of Interaction Risk Factors Associated with Suicidal Behaviors in Children with ADHD. Strength and direction of associations between predictors (psychiatric disorders and interactions with ADHD and Sex) and suicidal behaviors. Z-values derived from logistic regression analysis.

**Supplemental Table 1.** Demographic characteristics of the study cohort. descriptive statistics for the ABCD Study cohort, including total participants, sex distribution, mean age in months, and standard deviation of age.

| Demographic Metric | Count |
| --- | --- |
| Total Participants | 10,862 |
| Sex: Female | 5,198 |
| Sex: Male | 5,664 |
| Mean Age (months) | 118.9 |
| SD Age (months) | 7.5 |
| Ancestry: European (EUR) | 6,551 |
| Ancestry: African (AFR) | 2,107 |
| Ancestry: American (AMR) | 2,074 |
| Ancestry: South Asian (SAS) | 75 |
| Ancestry: East Asian (EAS) | 55 |
| Total ADHD Cases | 1,751 |
| ADHD: Male | 1,192 |
| ADHD: Female | 559 |
| Total ADHD Controls | 9,111 |
| Controls: Male | 4,472 |
| Controls: Female | 4,639 |

**Supplemental Table 2.** Suicide-related variables used in this study. Operational definitions and primary literature sources for the suicidal behavior variables included in the analysis.

| Variable | Definition | Citation(s) |
| --- | --- | --- |
| Active Suicidal Ideation | Desire to take action to kill oneself. Subdivided into: | Liu, Bettis, & Burke (2020) |
| – Active Intent | Thoughts that include a specific intention to die. |  |
| – Active Methods | Thoughts involving specific methods for suicide. |  |
| – Active Non-Specific | Desire to die without specific method or plan. |  |
| – Active Plan | Thoughts including a clear plan to carry out suicide. |  |
| Passive Suicidal Ideation | Thoughts of death or dying without specific plans or intention to act. |  |
| Preparatory Actions Toward Suicide | Actions taken to prepare for a suicide attempt before any injury occurs (e.g., collecting pills, writing a note, giving away possessions). | U.S. Dept. of Veterans Affairs, 2024 |
| Self-Injurious Behavior (No Suicidal Intent) | Deliberate self-injury without intent to die (e.g., cutting, burning). | U.S. Dept. of Veterans Affairs, 2024 |
| Suicide Attempt (Actual) | A non-fatal, self-directed act with the intent to die, regardless of the outcome. | Burke et al., 2016; Crosby et al., n.d. |
| Aborted Suicide Attempt | When an individual begins to act on suicidal thoughts but stops themselves before completing the act. | Barber et al., 1998; Posner et al., 2011b |
| Interrupted Suicide Attempt | A suicide attempt that is stopped by someone or something else before it can be completed. | Burke et al., 2016 |

**Supplementary Table 3.** Psychiatric diagnosis variables used in the analysis. List of psychiatric diagnoses analyzed in the study, with diagnostic category, status, and corresponding ICD-10 codes.

| Category | Diagnosis | Status | ICD-10 Code |
| --- | --- | --- | --- |
| Depression | Major Depressive Disorder | Present | F32.x |
|  | Major Depressive Disorder, in Partial Remission | Current | F32.4 |
|  | Major Depressive Disorder | Past | F32.9 |
|  | Persistent Depressive Disorder (Dysthymia), In Partial Remission | Current | F34.1 |
|  | Persistent Depressive Disorder (Dysthymia) | Past | F34.1 |
|  | Persistent Depressive Disorder (Dysthymia) | Present | F34.1 |
|  | Unspecified Depressive Disorder | Current | F32.9 |
|  | Unspecified Depressive Disorder | Past | F32.9 |
| Anxiety | Generalized Anxiety Disorder | Past | F41.1 |
|  | Generalized Anxiety Disorder | Present | F41.1 |
|  | Social Anxiety Disorder | Past | F40.10 |
|  | Social Anxiety Disorder | Present | F40.10 |
| Bipolar | Bipolar I Disorder, current episode depressed | Current | F31.3x |
|  | Bipolar I Disorder, current episode manic | Current | F31.1x |
|  | Bipolar I Disorder, currently hypomanic | Current | F31.02.4 |
|  | Bipolar I Disorder, most recent past episode depressed | Past | F31.1.3x |
|  | Bipolar I Disorder, most recent past episode manic | Past | F31.1x |
|  | Bipolar II Disorder, currently depressed | Current | F31.81 |
|  | Bipolar II Disorder, currently hypomanic | Current | F31.81 |
|  | Bipolar II Disorder, most recent past hypomanic | Past | F31.81 |
|  | Unspecified Bipolar and Related Disorder | Current | F31.9 |
|  | Unspecified Bipolar and Related Disorder | Past | F31.9 |

